# Striatal dopamine synthesis in schizophrenia decreases from psychosis to psychotic remission

**DOI:** 10.64898/2026.04.20.26351256

**Authors:** Julia Schulz, Melissa Thalhammer, Moritz Bonhoeffer, Viktor Neumaier, Franziska Knolle, Elisabeth F. Sterner, Qiongyu Yan, Rebecca Hippen, Stefan Leucht, Josef Priller, Wolfgang A. Weber, Yifan Mayr, Igor Yakushev, Christian Sorg, Felix Brandl

**Author notes:** **Corresponding author:** Felix Brandl, Department of Psychiatry and Psychotherapy, School of Medicine and Health, TUM Klinikum Rechts der Isar, Technical University of Munich, Ismaninger Str. 22, 81675 Munich, Germany.

## Abstract

Schizophrenia frequently follows a chronic relapsing-remitting course, comprising alternating episodes with and without psychotic symptoms (hereafter: psychosis and psychotic remission). One potential neurobiological correlate of this course is aberrant dopamine synthesis and storage (DSS) in the striatum, which can be estimated by ^18^F-DOPA positron emission tomography (PET). We hypothesised that striatal DSS in patients with schizophrenia decreases from psychosis to psychotic remission, with lower striatal DSS in patients during psychotic remission compared to healthy subjects. Additionally, we explored whether striatal DSS is associated with psychotic relapse after remission.

^18^F-DOPA PET scans and clinical assessments were conducted in 28 patients with schizophrenia at two timepoints, first during psychosis and second during early psychotic remission 6 weeks to 12 months after the first timepoint, as well as in 21 healthy controls, assessed twice in a comparable time interval. The averaged influx constant k_i_^cer^ as proxy for DSS was calculated for striatal subregions (i.e., nucleus accumbens, caudate, and putamen) using voxel-wise Patlak modelling with a cerebellar reference region. Mixed-effects models and post hoc analyses were used to test for longitudinal changes in k_i_^cer^ and cross-sectional group differences. An exploratory clinical follow-up 12 months after the second scan was conducted to assess psychotic relapse, and post hoc ANCOVAs were used to test for differences in k_i_^cer^ at each session between relapsing and non-relapsing patients.

K_i_^cer^ in both caudate and nucleus accumbens significantly changed from psychosis to psychotic remission compared to healthy controls, with a significant longitudinal decrease of caudate k_i_^cer^ in patients. Furthermore, k_i_^cer^ in both caudate and accumbens was significantly lower in patients during early psychotic remission compared to controls. At the exploratory clinical follow-up, 32% of patients had experienced a psychotic relapse; they showed higher caudate k_i_^cer^ compared to non-relapsing patients during psychosis, with no difference during psychotic remission.

These findings provide evidence for the link between striatal, particularly caudate, DSS and the relapsing-remitting course of psychotic symptoms in schizophrenia, with lower caudate DSS during early psychotic remission. Data suggest altered striatal dopamine synthesis together with impaired DSS dynamics along the course of psychotic symptoms in schizophrenia.

## Introduction

Schizophrenia is a debilitating psychiatric disorder that affects about 1% of the population worldwide.^1^ Its symptoms are heterogeneous and commonly categorised into three clusters: psychotic/positive (e.g. hallucinations, delusions), negative (e.g. anhedonia, social withdrawal) and cognitive (e.g. deficits in attention and executive function).^2,3^ Schizophrenia frequently follows a chronic relapsing-remitting course, comprising phases with and without psychotic symptoms in alternation, while negative and cognitive symptoms often persist rather continuously.^4^ Already within 12 months after remission of psychotic symptoms, there is a 25-30% risk of relapse of psychotic symptoms, even when antipsychotic treatment is continued.^4–6^ Despite sometimes conflicting evidence,^7^ there are hints that early relapse appears to predict poor long-term outcome.^8^ For this and other reasons, antipsychotic treatment maintenance for at least one year after psychotic remission is often recommended in clinical practice, despite changing and sometimes inconsistent guidelines.^9,10^ However, the underlying pathophysiological mechanisms of the relapsing-remitting course of psychotic symptoms in schizophrenia, particularly in the early phases of psychotic remission, remain poorly understood.

One promising neurobiological correlate of psychotic remission and relapse in schizophrenia is aberrant dopamine transmission in the striatum,^11–19^ particularly altered dopamine synthesis and storage (DSS), which can be estimated in-vivo using ^18^F-DOPA PET. Several meta-analyses have suggested increased striatal DSS, particularly in the dorsal striatum, as one of the most consistent in-vivo molecular imaging findings in schizophrenia.^13,15^ However, several limitations and open questions apply to these findings.

First, knowledge about the nature and trajectory of striatal DSS changes across relapse-remission states or cycles remains incomplete. For example, increased striatal DSS might not be specific to psychotic phases in schizophrenia, having also been observed in prodromal stages of schizophrenia, i.e. prior to the onset of psychosis,^20^ and other neurological and psychiatric disorders with psychotic symptoms, such as bipolar disorder and temporal lobe epilepsy.^11,21^ Furthermore, dopaminergic dysfunction in schizophrenia appears to be heterogeneous, since not all studies have reported increased striatal DSS in patients with active psychotic symptoms and some findings suggest lower striatal DSS in chronic patients with only mild psychotic symptoms.^19,22–25^ A previous ^18^F-DOPA PET study by our group, which investigated patients with schizophrenia during explicitly operationalised psychotic remission following Andreasen criteria^26^, observed decreased striatal DSS.^14^ Thus, these cross-sectional findings raise the possibility that striatal DSS may fluctuate across disease phases in schizophrenia, possibly being increased during acute psychosis and decreased during psychotic remission. Striatal DSS may therefore serve as a state-dependent marker of psychosis or its remission in schizophrenia. To further clarify this trajectory and its clinical relevance, longitudinal studies are necessary. However, apart from a single-patient report that observed increasing striatal DSS from remission to psychotic relapse 10 months later,^27^ direct longitudinal evidence tracking striatal DSS from psychosis to psychotic remission in patients with schizophrenia is currently lacking.

Second, adding to the scarcity of studies investigating striatal DSS during psychotic remission in schizophrenia, there are no studies of striatal DSS during early psychotic remission, i.e. remission lasting approximately 6–10 weeks, up to 12 months after measuring DSS during a psychotic episode. Since early periods of psychotic remission might be of particular clinical interest (see above), understanding dopamine function during early psychotic remission may have ramifications for treatment guidelines, including recommendations for the duration of antipsychotic treatment. However, previous ^18^F-DOPA PET studies in schizophrenia either did not define psychotic remission explicitly, i.e. not applying standardised remission criteria such as those by Andreasen et al.^25^ (e.g. reference ^22^), or included patients with widely varying durations of psychotic remission, often in the range of years.^14^ In this case, chronic dopaminergic deficits may obscure dynamic changes in dopaminergic transmission that occur shortly after psychotic remission. Therefore, ^18^F-DOPA PET studies in patients with schizophrenia in early psychotic remission are needed.

Based on these considerations, we conducted a longitudinal ^18^F-DOPA PET study (Fig. 1) in 28 patients and 21 matched controls. We hypothesised, first, a decreasing trajectory of striatal DSS in patients with schizophrenia from psychosis to psychotic remission, with – relative to healthy controls – increased striatal DSS during acute psychosis and decreased striatal DSS during psychotic remission. Second, performing a cross-sectional group comparison within the same study design, we hypothesised reduced striatal DSS in patients with schizophrenia during explicitly early psychotic remission (six weeks to 12 months after the session during psychosis) compared to controls. To test these hypotheses, we conducted two sessions of ^18^F-DOPA PET, accompanied by clinical and cognitive assessments, in patients with schizophrenia: the first during a psychotic episode and the second after psychotic remission, reached after at least six weeks and at most 12 months after the first measurement, thus corresponding to *early* psychotic remission. To enhance diagnostic specificity and distinguish schizophrenia from first-episode psychosis, we only included patients with at least one prior psychotic episode. A matched healthy control group was measured twice at comparable time intervals.

**Figure 1.**
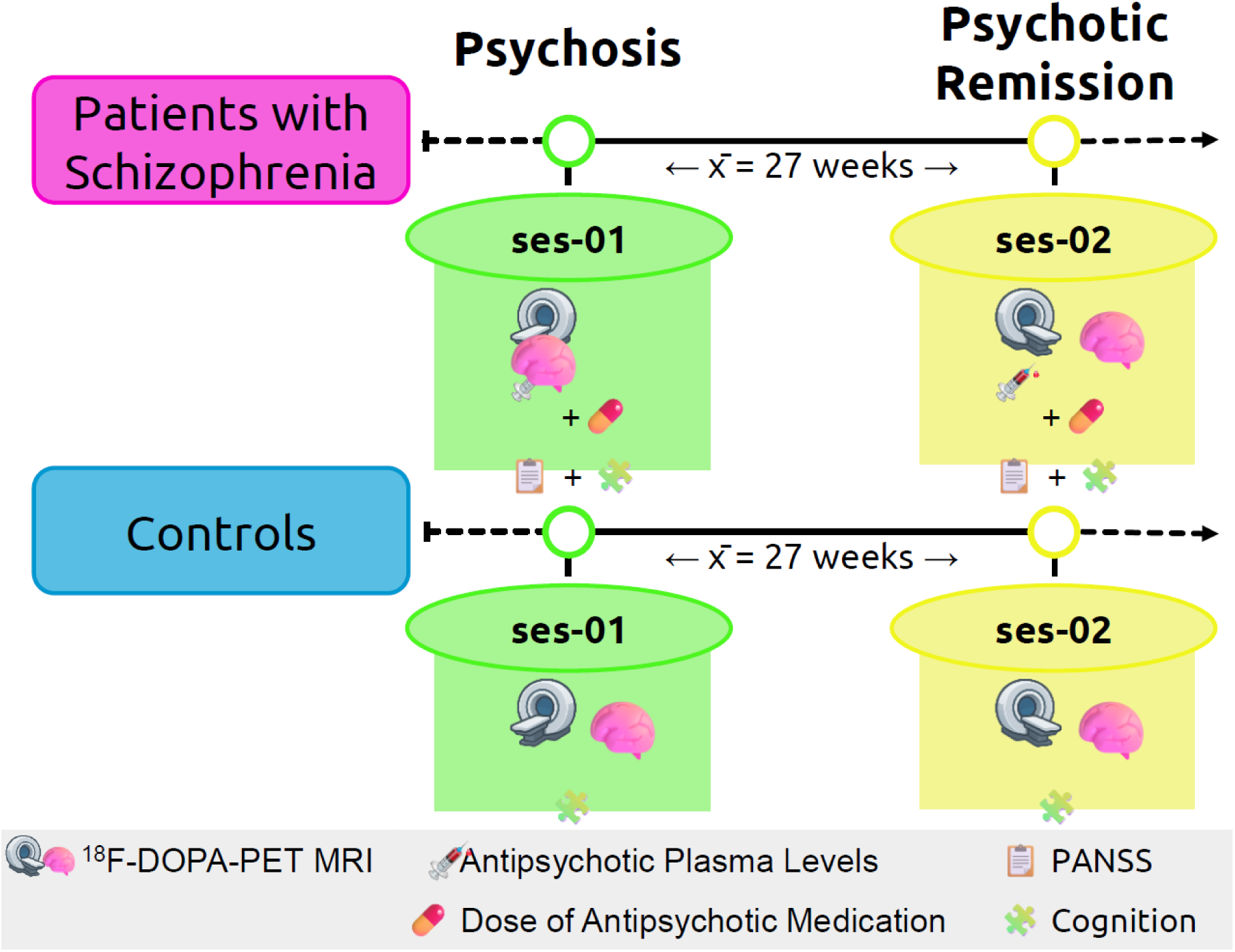
Longitudinal study design – from psychosis to psychotic remission. Patients with schizophrenia underwent two PET-MRI scans: the first during acute psychosis and the second during psychotic remission (mean between-scan interval = 27 weeks). In addition to imaging, plasma levels of antipsychotic medication, oral antipsychotic doses, clinical symptoms (PANSS) and cognitive performance were assessed. Healthy controls were also scanned twice in a comparable between-scan interval and completed cognitive testing. ses-01 session 1, ses-02 session 2, PANSS positive and negative syndrome scale.

As an additional exploratory investigation, we investigated whether striatal DSS at both timepoints (i.e., psychosis and psychotic remission) was associated with psychotic relapse within 12 months after remission. This is a relevant issue since, among other risk factors, discontinuation of antipsychotic medication, which prominently acts on the dopamine system, is a major predictor of relapse.^28,29^ Again, the first 12 months after psychotic remission might be of particular interest due to the common clinical practice of recommending continuing antipsychotic treatment for at least 12 months after psychotic remission. However, prospective evidence investigating the relevance of preceding striatal DSS changes for (early) psychotic relapse in schizophrenia is lacking. To address this question, we conducted an exploratory clinical follow-up in patients 12 months after the second ^18^F-DOPA PET measurement to determine the occurrence of psychotic relapse within this period. Relapse was then linked with striatal DSS by studying post hoc DSS differences between relapsing and non-relapsing patients during psychosis and during psychotic remission.

## Materials and methods

### Participants

The study was approved by the ethics review board of the TUM School of Medicine and Health, Technical University of Munich, Germany (approval No. 338-20 S/KH), and by the German Federal Office for Radiation Protection (“Bundesamt für Strahlenschutz”). All participants provided written informed consent before inclusion. General inclusion criteria were age between 18 and 65 years, no pregnancy, and ability to consent.

Patients with schizophrenia were recruited from the Department of Psychiatry of the University Hospital of Technical University of Munich (TUM Klinikum Rechts der Isar). All patients met DSM-5 criteria for schizophrenia,^30^ as examined by an experienced psychiatrist (F.B.), and were assessed using the Positive and Negative Syndrome Scale (PANSS).^31^ To ensure diagnostic certainty, all patients had experienced at least one prior psychotic episode that was not attributed to substance use. Exclusion criteria included substance abuse (except for nicotine) at least 12 months before and during the study, neurodevelopmental disorders, current or past neurological or neurosurgical diseases and severe systemic diseases. No restrictions were applied regarding the type of antipsychotic medication or its receptor affinity. Benzodiazepines were not permitted prior to the scan. No further restrictions were applied regarding adjuvant medication. At the time of the first session, patients had to be in an acute psychotic episode for at least two weeks, defined in line with previous literature as at least moderate severity (≥4) in at least one of the PANSS items “delusions” (P1), “hallucinatory behavior” (P3) or “suspiciousness/persecution” (P6).^21,31,32^ After the first session, patients were monitored regularly (every four weeks) by an experienced psychiatrist (F.B.) to track symptom remission and ensure treatment adherence.

For the second session, patients had to be in remission of psychotic symptoms for at least six weeks following criteria by Andreasen et al.^26^, i.e., the PANSS items “delusions” (P1), “conceptual disorganization” (P2), “hallucinatory behavior” (P3), “suspiciousness/persecution” (P6), “mannerisms/posturing” (G5) and “unusual thought content” (G9) all had to be ≤3. No remission was required for negative or general symptoms. Psychotic remission had to be achieved within 12 months after the first session; otherwise, patients were excluded from the study.

Additionally, healthy subjects comparable in age and sex were recruited from the Munich area. They also underwent two measurement sessions in a comparable time interval. Additional exclusion criteria for the healthy controls comprised any current or past psychiatric disorder (assessed via the Structured Clinical Interview for DSM-5), use of psychotropic medication and family history of psychosis.

### Clinical assessments and medication

At each session, patients’ symptom severity was assessed by the same experienced psychiatrist (F.B.) using PANSS.^31^ Cognitive performance was evaluated in all participants using the Trail Making Test Part B (TMT-B) and the Symbol Coding Task (SCT). TMT-B measures visual-motor processing speed, executive function, cognitive flexibility and working memory, and is reported as completion time in seconds.^33^ SCT evaluates processing speed and attention and is reported as the number of correct symbol-digit matches within 90 seconds.^34^

To ensure medication adherence and for statistical control purposes, both plasma levels and oral doses of antipsychotic medication were measured and quantified. Oral medication dose was standardised as chlorpromazine (CPZ) equivalents using the R package ‘chlorpromazineR’ (https://github.com/ropensci/chlorpromazineR).^35^ Antipsychotic plasma levels were measured before each scan and categorised with regard to their clinical reference range:^36^ 0 = not detectable; 1 = subtherapeutic (below the reference range); 2 = within the therapeutic range; 3 = supratherapeutic (above the reference range). When a patient took multiple antipsychotics, these scores were summed.

### 18F-DOPA PET data acquisition

All participants underwent scanning at the university hospital of Technical University of Munich, Germany, using a hybrid whole-body PET/MRI system (mMR Biograph, Siemens-Healthineers, Erlangen, Germany) with a 32-channel phase-array head coil. Our PET protocol followed previous studies.^14,37^ Participants were positioned in the scanner with the orbitomeatal line parallel aligned to the transaxial plane. ^18^F-DOPA (mean administered dose: 153.99 ± 9.36 MBq) was injected intravenously as a bolus at the start of scanning, followed by a saline flush. PET data were acquired for 70 min. Reconstruction was performed using the Siemens syngo MR E11 offline PET reconstruction toolkit, incorporating attenuation, scatter, randoms and radioactive decay correction. Ordered-subsets expectation maximization (OSEM) was applied with 24 subsets and three iterations, with a 3 mm Gaussian post-reconstruction filter. Serial PET data with a voxel size of 1.04 x 1.04 x 2.03 mm³ were divided into 30 frames: 1 × 30 s, 10 × 15 s, 3 × 20 s, 2 × 60 s, 2 × 120 s and 12 × 300 s.

Structural MRI data using a T1-weighted MPRAGE sequence (TR/TE/flip angle: 2300 ms/2.98 ms/9°; FoV: 256 mm; matrix size: 160×240×256; voxel-size: 1×1×1 mm³) were acquired simultaneously for subsequent coregistration.

### 18F-DOPA PET data processing

PET data were preprocessed using a pipeline developed in-house (PET-ki-proc), publicly available at https://github.com/juliasbrain/PET_ki_proc (Supplementary Fig. 1), following references ^13,36^. All analyses were performed in Python 3.11.5, using FSL version 6.0.7.4. A detailed description of PET data preprocessing is given in the GitHub repository and Supplementary Methods.

Motion correction using FSL *mcflirt* was applied to the attenuation-corrected dynamic PET data from 5 to 70 minutes post-injection, using the last frame as reference.^37^ The motion-corrected data were then denoised using Chambolle’s total variation denoising algorithm implemented via the scikit-image Python package. Each participant’s T1-weighted image was coregistered to the last PET frame using linear transformation (FSL *flirt*) and subsequently normalised to MNI-152 standard space using non-linear transformation (FSL *flirt* and *fnirt*). 3D regions of interest (ROIs) comprised nucleus accumbens, caudate and putamen from a probabilistic anatomical atlas^38^ and a cerebellar reference region from a probabilistic cerebellum atlas^39^. ROIs were transformed into native PET space using the inverse warp of the normalisation step. Then, the influx constant k_i_^cer^, interpreted as DSS, was calculated voxel-wise using Gjedde–Patlak linear graphical analysis with the cerebellum as reference region.^40^ In line with previous studies,^14,37^ the linear fit was applied to PET frames between 20 and 60 minutes post-injection. Finally, k_i_^cer^ values were averaged for each striatal subregion.

### Statistical analyses

Statistical analyses were carried out using Python 3.11.5 and R 4.2.0. To test assumptions for parametric testing, Shapiro–Wilk tests for normality and Levene’s tests for homoscedasticity were used. Demographic and clinical variables (e.g. symptom scores and medication) were compared between groups using independent two-sample t-tests, Mann–Whitney U-tests, chi-square tests, linear mixed-effects models, or generalised linear mixed models with a logistic link for binary dependent variables.

To examine longitudinal changes in striatal DSS from psychosis to psychotic remission, linear mixed-effects models were used, accounting for repeated measures within subjects and accounting for dropouts. For each striatal subregion, the dependent variable was mean k_i_^cer^, with group, session and their interaction (group x session) as fixed effects, and subject as random effect. For post hoc pairwise comparisons between groups and sessions, one-sided t-tests were performed on k_i_^cer^ values based on *a priori* hypotheses of increased k_i_^cer^ during psychosis and decreased k_i_^cer^ during psychotic remission.

### Reliability and control analyses

To test for the reliability of our measurements and to control for the influence of methodological and clinical factors and medication, several control analyses were performed. First, the reliability of k_i_^cer^ measures in healthy controls was assessed by comparing k_i_^cer^ values across striatum, midbrain and cortical grey matter as well as by assessing the test-retest reliability using intraclass correlation coefficients. Second, we tested for the influence of imaging-based methodological variables on results, namely the reference frame for motion correction and the size of striatum and cerebellum ROIs. Third, the influence of demographic and clinical variables as well as medication on results was evaluated, namely age, sex, smoking status, between-scan interval, CPZ equivalents and antipsychotic plasma levels. Fourth, we investigated the effect of specific antipsychotic drugs on the results, namely clozapine, which might particularly influence DSS according to a previous study,^22^ aripiprazole, a partial dopamine receptor agonist possibly influencing DSS,^41^ and amisulpride, a rather selective D2 antagonist that could affect dopamine transmission in a particular way.^42,43^

### Exploratory clinical follow-up linking psychotic relapse within 12 months and preceding striatal DSS

As an exploratory analysis under clinical conditions, without specific control of treatment or confounding factors (e.g. medication adherence, psychotherapy or stressful life events), the potential relevance of striatal DSS at both timepoints (i.e., psychosis and psychotic remission) for a subsequent psychotic relapse was investigated. At a clinical follow-up in patients 12 months after session 2, the relapse of psychotic symptoms since the second session was assessed and the current antipsychotic medication was recorded. ANCOVAs were used to assess differences in striatal k_i_^cer^ between relapsing and non-relapsing patients during psychosis (session 1) and psychotic remission (session 2), controlling for the between-scan interval, which significantly differed between both patient subgroups.

## Results

### Demographics and clinical characteristics

Twenty-eight patients (mean age: 35.63 ± 9.56, range: 21-61 years, female/male: 9/19) and 21 controls (mean age: 33.46 ± 12.31, range: 22-62 years, female/male: 12/9) were included in the study and completed the first session. Of these participants, 23 patients and 20 controls also completed the second session. One patient did not remit within the predefined 12-month period after the first session. The remaining dropouts (four patients and one control) were due to other reasons, such as relocation or loss of contact.

Demographic and clinical variables are summarized in Table 1. Patients and controls did not differ regarding age (ses-01: *t* = -0.7, *P* = 0.5), sex (χ^2^ = 2.1, *P* = 0.14) or between-scan interval (*t* = 0.19, *P* = 0.85). Nicotine use was more prevalent in patients at both sessions (linear mixed-effects model: group χ^2^ = 15.8, *P* < 0.001; session: χ^2^ = 0.1, *P* = 0.97; post hoc comparisons: ses-01: *z* = -3.4, *P* < 0.001; ses-02: *z* = -3.3, *P* = 0.001). The injected radioactivity did not differ significantly between groups or across sessions (linear mixed-effects model: group *F*(1,48) = 0.3, *P* = 0.58; session: *F*(1,40) = 0.7, *P* = 0.41).

**Table 1.** Demographic and clinical variables. Data are presented as means and standard deviations. Demographic variables were compared between groups using independent two-sample t-tests (1), Mann-Whitney U tests (2), chi-square tests (3), linear mixed-effects models (4), or generalized linear mixed models with a logistic link (5) for binary dependent variables. Antipsychotic plasma levels were categorized with respect to their clinical reference range: 0=not detectable, 1=below the reference range, 2=within the therapeutic range, 3=above the reference range; when a patient received multiple antipsychotics, these scores were summed up. *GLMM generalized linear mixed-effects model, HC healthy controls, LMM linear mixed-effects model, PANSS positive and negative syndrome scale, SCT symbol coding task, SCZ patients with schizophrenia, TMT-B Trail Making Test part B*.

|  | Session | HC | SCZ | Between-group comparison | Between-session comparison |
| --- | --- | --- | --- | --- | --- |
| n | 1 | 21 | 28 |  |  |
|  | 2 | 20 | 23 |  |  |
| Between-scan interval [d] | | 189.0 (± 79.84) | 191.86 (± 97.61) | $t = 1.9, P = 0.85^1$ | |
| Age [y] | 1 | 33.46 (± 12.31) | 35.63 (± 9.56) | $t = -0.7, P = 0.5^1$ | |
| | 2 | 34.48 (± 12.36) | 37.13 (± 10.01) | $t = -0.8, P = 0.5^1$ | |
| Sex (f/m) | 1 | 12 / 9 | 9 / 19 | $\chi^2 = 2.1, P = 0.14^3$ | |
| | 2 | 11 / 9 | 8 / 15 | $\chi^2 = 1.04, P = 0.31^3$ | |
| Smoking (no/yes) | 1 | 20 / 1 | 11 / 17 | $\chi^2 = 15.8, P < 0.001^5$ | $\chi^2 = 0.1, P = 0.97^5$ |
|  | 2 | 19 / 1 | 9 / 14 |  |  |
| Injected $^{18}\text{F}$-DOPA activity [Mbq] | 1 | 154.57 (± 7.25) | 154.82 (± 8.47) | $F = 0.3, P = 0.58^4$ | $F = 0.7, P = 0.41^4$ |
| | 2 | 151.95 (± 11.81) | 154.22 (± 9.24) | | |
| Illness Duration [y] | 1 | | 12.07 (± 8.83) | | |
| | 2 | | 13.78 (± 8.93) | | |
| Chlorpromazine equivalents [mg/d] | 1 | | 470.57 (± 384.74) | | $U = 322, P = 0.99^2$ |
| | 2 | | 441.54 (± 311.39) | | |
| Antipsychotic plasma levels [a.u.] | 1 | | 3.18 (± 3.04) | | $U = 266.0, P = 0.28^2$ |
| | 2 | | 2.22 (± 1.28) | | |
| PANSS total [a.u.] | 1 | | 83.71 (± 14.58) | | $t = -8.8, P < 0.001^1$ |
| | 2 | | 52.87 (± 9.27) | | |
| PANSS positive [a.u.] | 1 | | 22.32 (± 3.39) | | $t = -11.5, P < 0.001^1$ |
| | 2 | | 12.39 (± 2.61) | | |
| PANSS negative [a.u.] | 1 | | 21.89 (± 5.93) | | $U = 93.5, P < 0.001^2$ |
| | 2 | | 14.96 (± 4.13) | | |
| TMT-B [sec] | 1 | 52.52 (± 21.30) | 102.04 (± 52.23) | $F = 23.3, P < 0.001^4$ | $F = 3.5, P = 0.07^4$ |
| | 2 | 51.54 (± 18.61) | 86.83 (± 37.69) | | |
| SCT [a.u.] | 1 | 65.00 (± 9.96) | 41.21 (± 9.20) | $F = 81.2, P < 0.001^4$ | $F = 2.6, P = 0.11^4$ |
| | 2 | 66.80 (± 11.26) | 42.26 (± 12.94) | | |

In patients, psychotic symptoms, as measured by PANSS positive (PANSS+), were significantly higher during psychosis than during psychotic remission (PANSS+: ses-01 = 22.32 ± 3.39, ses-02 = 12.39 ± 2.61; *t* = -11.5, *P* < 0.001). At the individual level, all patients had lower PANSS positive scores in psychotic remission compared to psychosis, indicating a consistent decrease in positive symptoms during remission (Fig. 2). Negative and total symptoms (PANSS negative and total) also improved on average during psychotic remission, while cognitive impairments (lower cognitive performance in TMT-B and SCT) did not change along the course (see Supplementary Results, Supplementary Fig. 2, 3).

**Figure 2.**
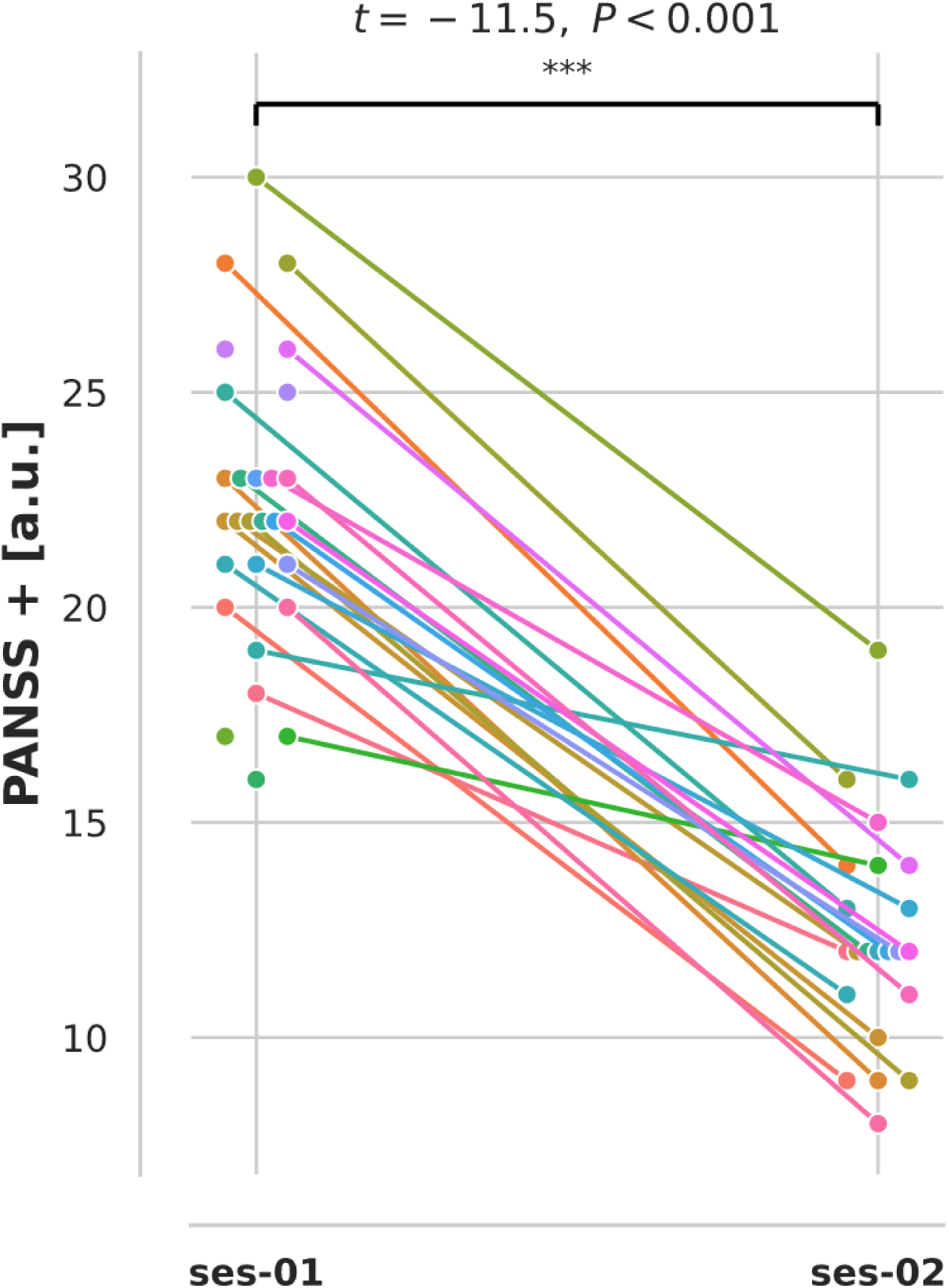
Individual decrease in psychotic symptoms (quantified by PANSS positive) in patients with schizophrenia from psychosis (ses-01) to psychotic remission (ses-02). Two-sample t-test of PANSS positive during psychosis (ses-01) and psychotic remission (ses-02): *t* = -11.5, *P* < 0.001. \**P* ≤ 0.05, \*\**P* ≤ 0.01, \*\*\**P* ≤ 0.001. ses-01 session 1, ses-02 session 2, PANSS positive and negative syndrome scale.

Oral antipsychotic dose (CPZ equivalents, ses-01 = 470.57 ± 384.74 mg/d, ses-02 = 441.54 ± 311.39 mg/d, *U* = 322, *P* = 0.99) and antipsychotic plasma levels (ses-01 = 3.18 ± 3.04, ses-02 = 2.22 ± 1.28, *U* = 266, *P* = 0.28) did not differ significantly between sessions in the patient group. A list of antipsychotics is shown in the Supplement (Supplementary Fig. 4C).

### Decreasing caudate k_i_^cer^ from psychosis to psychotic remission, with reduced k_i_^cer^ in remission

Linear mixed-effects models showed a significant group x session interaction on k_i_^cer^ in nucleus accumbens (*F*(1,40) = 4.4, *P* = 0.041) and caudate (*F*(1,40) = 4.8, *P* = 0.034), indicating that DSS changed from psychosis to psychotic remission in patients compared to controls (Fig. 3). No main effects of group or session were observed. In the putamen, no significant effects were detected for group, session or their interaction (group x session: *F*(1,40) = 0.1, *P* = 0.75). Based on these findings, subsequent analyses focused on the nucleus accumbens and caudate.

**Figure 3.**
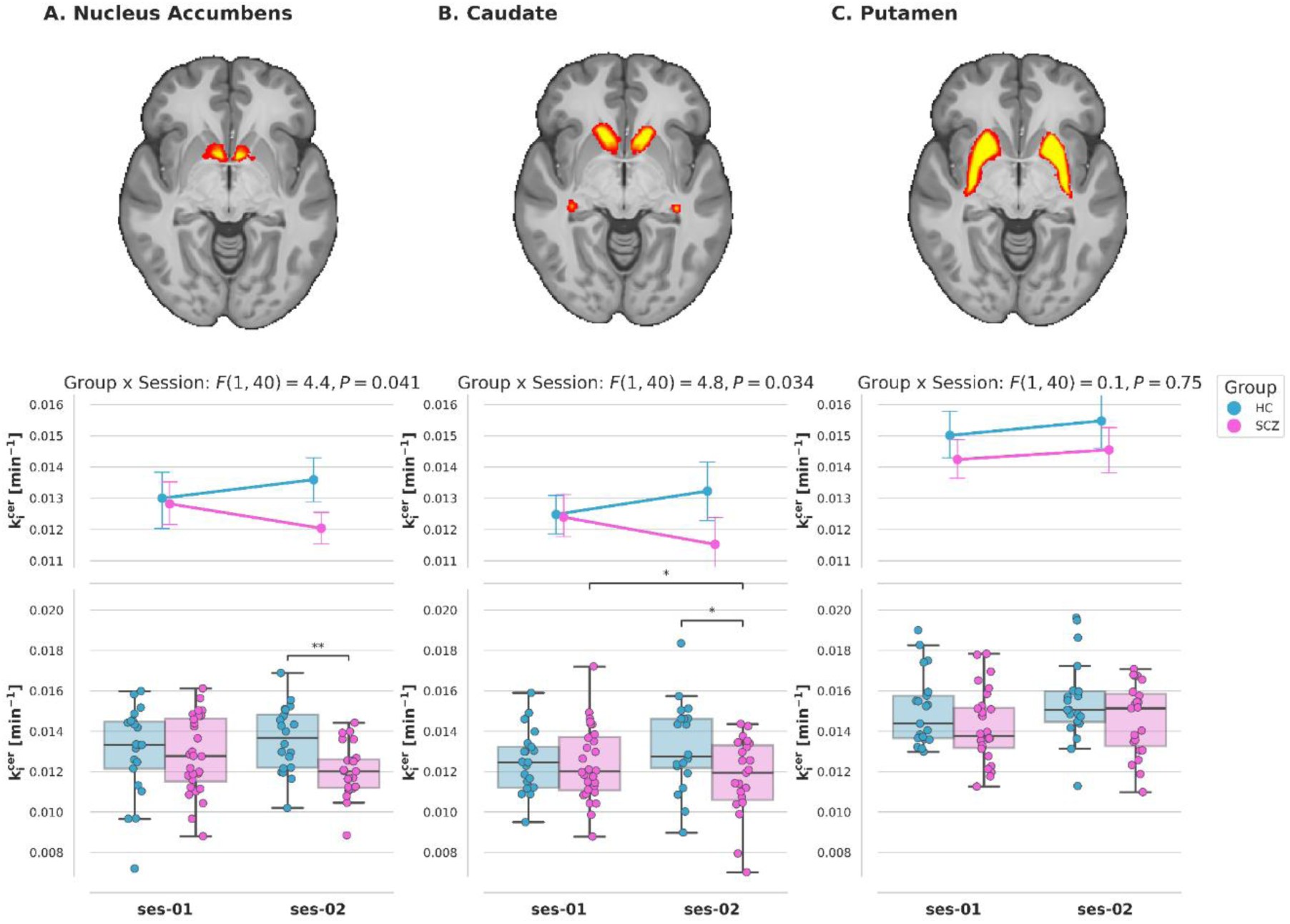
Decrease to lower striatal dopamine synthesis and storage from psychosis to psychotic remission in schizophrenia. A linear mixed-effects model was used to investigate the effects of group, session, and their interaction on k_i_^cer^ in nucleus accumbens (**A**), caudate (**B**), and putamen (**C**). There was a significant group x session interaction in nucleus accumbens (*F*(1,40) = 4.4, *P* = 0.041) and caudate (*F*(1,40) = 4.8, *P* = 0.034. (**A**) For the nucleus accumbens, post hoc tests showed a significantly lower k_i_^cer^ in patients during psychotic remission compared to controls at session 2 (*t* = 2.4, *P* = 0.01). (**B**) For the caudate, post hoc tests showed higher k_i_^cer^ in patients during psychosis compared to psychotic remission (*t* = 1.8, *P* = 0.043), and lower k_i_^cer^ in patients during psychotic remission compared to controls at session 2 (*t* = 2.4, *P* = 0.02). \**P* ≤ 0.05, \*\**P* ≤ 0.01, \*\*\**P* ≤ 0.001. HC healthy controls, k_i_^cer^ influx constant of ^18^F-DOPA relative to a cerebellar reference region, ses-01 session 1 (in patients: psychosis), ses-02 session 2 (in patients: psychotic remission), SCZ patients with schizophrenia.

Pairwise post hoc comparisons showed a significant longitudinal decrease in k_i_^cer^ in patients from psychosis to psychotic remission in the caudate (*t* = 1.8, *P* = 0.043) but not in the nucleus accumbens (*t* = 1.5, *P* = 0.065). In healthy controls, no longitudinal changes were observed (Supplementary Material). Cross-sectionally, we found significantly decreased k_i_^cer^ in patients during psychotic remission compared to healthy controls at session 2 in both the nucleus accumbens (*t* = 2.4, *P* = 0.01) and the caudate (*t* = 2.1, *P* = 0.02), while no significant differences in k_i_^cer^ between patients during psychosis and healthy controls at session 1 were found in either nucleus accumbens (*t* = 0.01, *P* = 0.51) or caudate (*t* = 0.4, *P* = 0.67). An overview of all results is provided in Table 2.

**Table 2.**
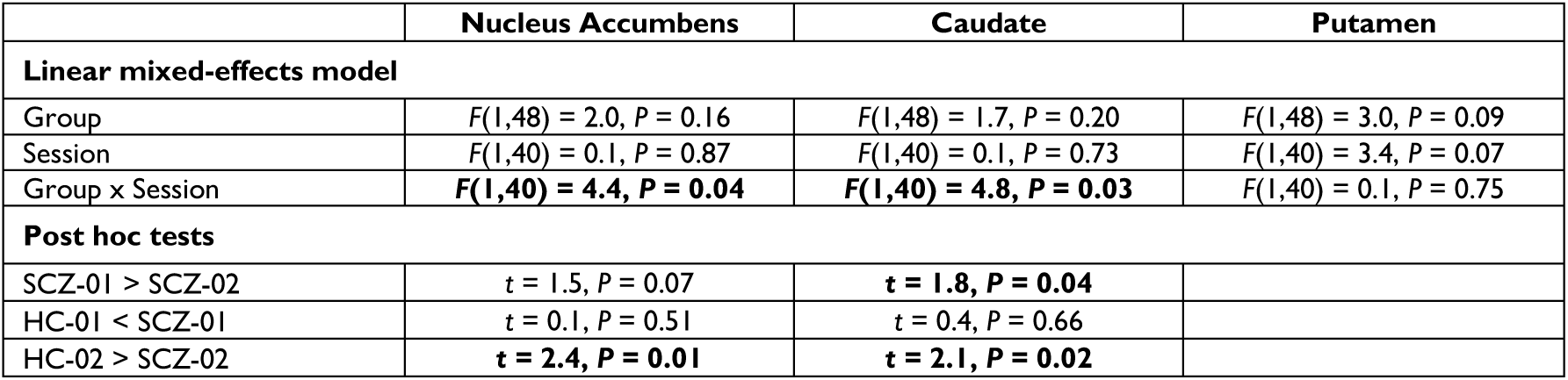
Longitudinal and cross-sectional changes in k_i_^cer^ from psychosis to psychotic remission – linear effects model analysis. The table shows main effects and post hoc comparisons (one-sided t-tests) for each striatal subregion. Significant effects are highlighted in bold font. *HC healthy controls; SCZ patients with schizophrenia; 01 session 1 (in patients: psychosis); 02 session 2 (in patients: psychotic remission)*.

In summary, we found that caudate k_i_^cer^ decreased in patients with schizophrenia in the course from psychosis to psychotic remission, with lower values during psychotic remission. A similar pattern was observed for k_i_^cer^ in the nucleus accumbens, although the longitudinal decrease was not statistically significant in post hoc analyses.

### Results of the reliability and control analyses, including control for antipsychotic medication

To ensure that the observed alterations in striatal k_i_^cer^ in patients were based on a reliable measurement approach and were not influenced by methodological, demographic or clinical factors as well as current medication, we performed several control analyses (for details, see Supplementary Material: Control and reliability analyses). First, in line with previous studies,^13,43^ we found that k_i_^cer^ values in healthy controls were highest in the striatum, intermediate in the midbrain and lowest in cortical grey matter across both sessions.

Furthermore, and importantly, healthy controls’ k_i_^cer^ values showed sufficient test-retest reliability, demonstrating reliable k_i_^cer^ measurements. Second, regarding methodological variables of PET data analysis, the results were not influenced by the choice of the reference frame in motion correction or the size of striatum and cerebellum masks. Third, the main result of the group × session interaction in the nucleus accumbens and caudate was not influenced by age, sex, smoking, between-scan interval or antipsychotic medication (CPZ equivalents and plasma levels). Fourth, treatment with specific antipsychotics, i.e. clozapine, aripiprazole or amisulpride, did not significantly affect the results.

### Exploratory follow-up: link of caudate k_i_ ^cer^ during psychosis and psychotic relapse within 12 months

At the exploratory clinical follow-up 12 months after session 2 (psychotic remission), 7 out of 22 reached patients with longitudinal data had experienced a psychotic relapse, i.e. 32%, which is consistent with relapse rates reported in the literature.^5,6^ We found no differences between relapsing and non-relapsing patients regarding antipsychotic medication or psychotic symptoms during both psychosis and psychotic remission (detailed description of results in the Supplement). Interestingly, the between-scan interval was significantly longer in relapsing patients, which potentially hints at a slower initial recovery from psychosis.

Then, focusing on striatal DSS preceding psychotic relapse, we studied differences in k_i_^cer^ between relapsing and non-relapsing patients using ANCOVAs, controlling for the between-scan interval as covariate of no interest. Group comparisons were conducted for caudate and nucleus accumbens, since DSS in both regions had changed from psychosis to psychotic remission. During psychosis, we observed significantly higher k_i_^cer^ values in the caudate in patients who later relapsed compared with those who did not (*F*(1,19) = 4.4, *P* = 0.049) (Fig. 4). In contrast, no group differences were found in the caudate during psychotic remission (*F*(1,19) = 0.6, *P* = 0.45) or in the nucleus accumbens during psychosis (*F*(1,19) = 3.0, *P* = 0.10) or psychotic remission (*F*(1,19) = 1.5, *P* = 0.24; Fig. 4). Thus, caudate DSS during psychosis, but not during psychotic remission, appears to be relevant for subsequent psychotic relapse within 12 months.

**Figure 4.**
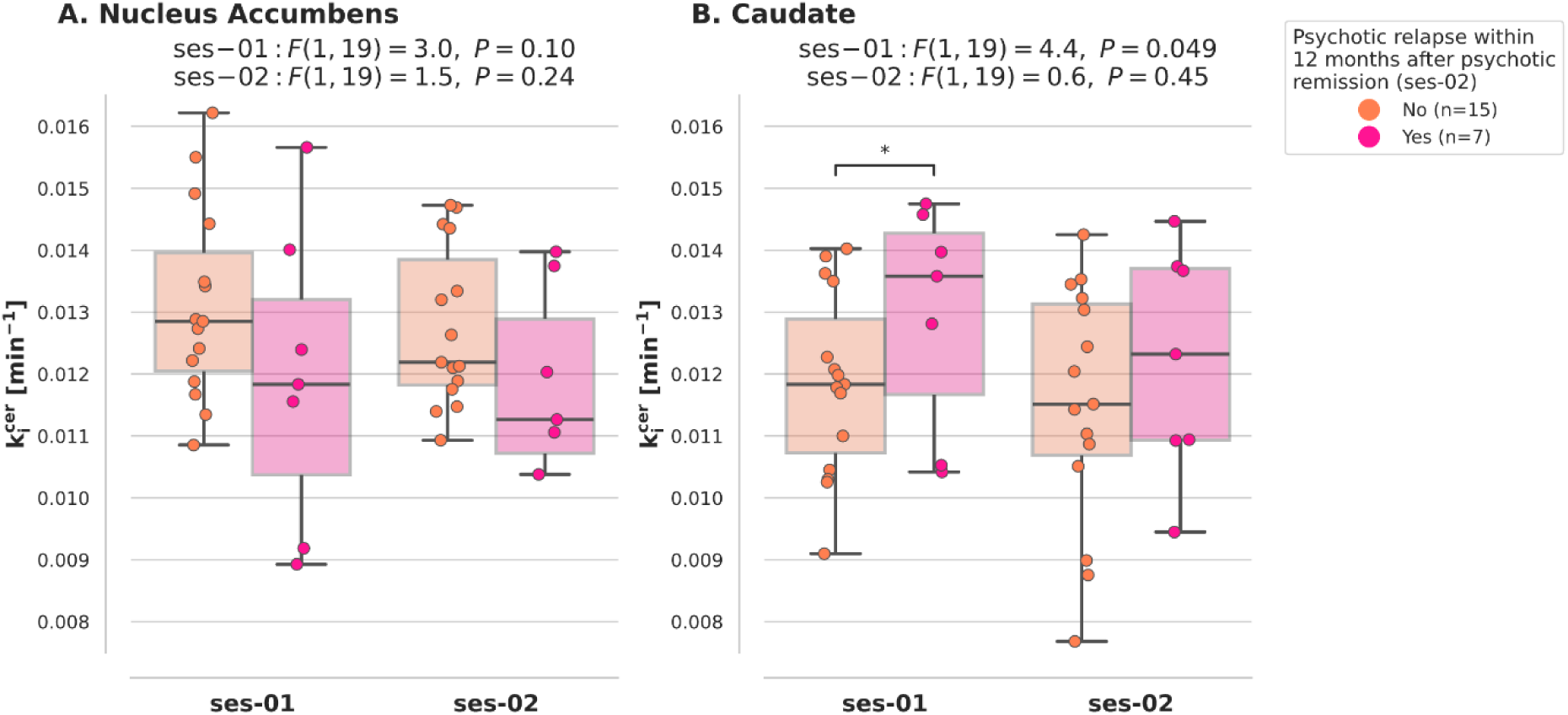
Clinical follow-up of psychotic relapse after remission: Higher caudate DSS during psychosis, but not psychotic remission, in patients with subsequent psychotic relapse. Group comparisons of k_i_^cer^ between patients who later relapsed (pink) and those who did not (orange) were performed using ANCOVA. (**A**) For the nucleus accumbens, no significant differences were observed in either session (ses-01: *F*(1,19) = 3.0, *P* = 0.10, ses-02: *F*(1,19) = 1.5, *P* = 0.24). (**B**) For the caudate, k_i_^cer^ was significantly higher in patients who relapsed compared to those who did not during session 1 (psychosis) (*F*(1,19) = 4.4, *P* = 0.049), but no difference was detected during session 2 (psychotic remission). \**P* ≤ 0.05, \*\**P* ≤ 0.01, \*\*\**P* ≤ 0.001. k_i_^cer^ influx constant of ^18^F-DOPA relative to a cerebellar reference region, ses-01 session 1 (psychosis), ses-02 session 2 (psychotic remission).

## Discussion

We conducted a longitudinal ^18^F-DOPA PET study in patients with schizophrenia and healthy controls to investigate changes in striatal subregions’ DSS in the course from acute psychosis to early psychotic remission. We observed that k_i_^cer^ in both caudate and nucleus accumbens significantly changed from psychosis to psychotic remission, with a significant longitudinal decrease in the caudate. Furthermore, in both caudate and nucleus accumbens, k_i_^cer^ was significantly lower in patients during early psychotic remission compared to controls, but did not differ during psychosis. These results remained significant when controlling for current antipsychotic medication overall and specific antipsychotics. Finally, in an exploratory clinical follow-up after 12 months, caudate k_i_^cer^ during psychosis, but not during psychotic remission, was linked with psychotic relapse after remission. To the best of our knowledge, these findings provide first-time evidence for the link between striatal, particularly caudate, DSS and the relapsing-remitting course of psychotic symptoms in schizophrenia, with lower caudate DSS during psychotic remission. Data suggest an impaired striatal dopamine synthesis (i.e. lower DSS during remission) together with impaired DSS dynamics (i.e. longitudinal DSS changes and link between DSS levels during psychosis and relapse risk) in schizophrenia.

### Decreasing caudate DSS from psychosis to psychotic remission – altered DSS dynamics in schizophrenia

Using linear mixed-effects models to analyse longitudinal ^18^F-DOPA PET data in striatal subregions, we found significant group x session interactions on k_i_^cer^ for both caudate and nucleus accumbens, but not the putamen (Fig. 3, Table 2). Post hoc tests showed a significant decrease in caudate k_i_^cer^ from psychosis to remission. These results were valid and reliable: we used established clinical criteria to define psychosis and psychotic remission in patients with schizophrenia,^21,26,32^ and reliability/control analyses found no substantial influence from methodological parameters (i.e. motion correction or mask size) or demographic and clinical variables including age, gender, smoking, between-scan interval and, particularly, current antipsychotic medication. Taken together, caudate and nucleus accumbens k_i_^cer^ significantly changed from psychosis to psychotic remission, with a significant longitudinal decrease in the caudate and the same trend (although not significant) in nucleus accumbens.

Interpreting our results from a signal-based point of view, our outcome measure k_i_^cer^ reflects the steady-state uptake rate of ^18^F-DOPA in the striatum relative to the cerebellum, based on the standard Gjedde-Patlak linear graphic analysis, and is generally interpreted as an estimate of presynaptic DSS.^45,46^ Regarding the temporal dynamics of DSS, there is limited prior research on DSS changes in the range of weeks. In contrast, there is ample research on changes of microscopical dopamine measures in the range of seconds or minutes: for example, ‘tonic’ (longer-term, referring to minutes) and ‘phasic’ (short-term, referring to seconds and less) dopamine signaling changes can be distinguished and are generally associated with reward-related learning and decision making, e.g. in references ^46,47^. Furthermore, striatal DSS changes over years have been observed during aging, with progressive DSS declines contributing to age-related cognitive changes such as cognitive flexibility.^49,50^ But there are few studies focusing on the temporal dynamics of (PET-based) DSS changes in the range of weeks and months: Previous longitudinal studies in healthy subjects over the span of both days and years have shown no significant DSS changes in the striatal subregions investigated here,^51,52^ while in schizophrenia, we only know of a single-patient report observing increasing striatal DSS from remission to psychotic relapse 10 months later.^27^ Thus, context and related underpinnings of the observed k_i_^cer^ changes during weeks/few months from psychosis to psychotic remission remain unclear. This lack of knowledge particularly concerns our incomplete understanding of the neurobiological mechanisms that control DSS and their temporal dynamics. Notwithstanding, we would like to mention some promising but speculative candidates for such control mechanisms: D2 autoreceptors on dopamine neurons,^53^ D2 receptors on striatal neurons,^54^ density of dopaminergic neuronal fibers^55^ and functional connectivity within cortico-striato-pallido-thalamo-cortical circuits.^14,56^ For example, D2 autoreceptors on dopamine neurons, which control dopamine transmission, appear to be regulated by various substances and reward-related plasticity,^53^ potentially contributing to slow changes in DSS. Future multimodal studies are needed for mechanistic clarification.

Moreover, regarding the regional specificity of our results, the focus of DSS changes on the caudate is generally consistent with the literature: although previous meta-analyses of caudate DSS in schizophrenia reported conflicting results,^12,13^ a more recent meta-analysis observed the most robust DSS changes in parts of the striatum called ‘associative’ due to connections with associative cortices, which greatly overlap with the caudate.^15^ For accumbens and putamen DSS in schizophrenia, evidence is mixed, with historical theories often focusing on the ventral striatum,^57^ while newer evidence points more to the dorsal striatum.^15^ Interestingly, a recent study reported that in schizophrenia, DSS in regions mostly overlapping with caudate and accumbens is specifically linked with mesencephalic neuromelanin, a measure of long-term dopamine metabolism.^58^ Future studies with possibly more fine-grained striatal subdivisions and integration with MRI-based measures are needed to clarify this issue.

Next, it is important to discuss the potential influence of antipsychotic medication on dopamine synthesis and its temporal dynamic: While drug effects have been shown on the microscopic level,^59^ results in PET studies remain inconsistent.^60–63^ Although, for pragmatic and ethical reasons, we did not investigate antipsychotic-naïve or antipsychotic-free patients and did not have a strict criterion that medication remain identical across both scan sessions, we statistically controlled for both oral antipsychotic dose (CPZ equivalents) and antipsychotic plasma levels and found no significant effect on the results. Furthermore, although medication changed from psychosis to psychotic remission in some patients, average CPZ equivalents and antipsychotic plasma levels did not significantly change (Table 1), implying largely stable antidopaminergic effects across both sessions. Neither did we find confounding influences from three antipsychotics with particularly representative properties, namely clozapine, which might distinctly influence DSS,^22^ aripiprazole, a partial dopamine receptor agonist possibly influencing DSS,^41^ and amisulpride, a rather selective D2 antagonist that could affect dopamine transmission in a particular way.^42,43^ Thus, we did not find any influence from current medication, implying that the observed DSS changes were indeed to the changes in psychotic symptoms. However, due to the lack of experimental control of medication and the lack of control of previous medication, our results should be interpreted with caution and followed up by studies employing stricter medication control or investigating antipsychotic-naïve/free patients.

### Lower caudate and nucleus accumbens DSS in early psychotic remission

The cross-sectional post hoc tests comparing patients during psychotic remission with healthy controls at session 2 showed decreased DSS in caudate and nucleus accumbens, but not in the putamen (Fig. 3, Table 2). Since the measurement in psychotic remission was conducted approximately 6-10 weeks after remission of psychotic symptoms, and maximally 12 months after the first measurement, patients were in *early* psychotic remission. Reliability/control analyses confirmed the robustness of these results regarding methodological and clinical variables and antipsychotic medication.

To the best of our knowledge, this is the first ^18^F-DOPA PET study in patients with schizophrenia during explicit early remission of psychotic symptoms. Our finding of decreased striatal DSS both confirms and extends our previous finding of decreased striatal DSS in patients with widely varying durations of psychotic remission, often in the range of years.^14^ Thus, striatal DSS is reduced already in early psychotic remission, and not only in chronic remission, where chronic dopaminergic deficits might dominate.

This finding raises several questions, for example: Does striatal DSS reduction during psychotic remission remain stable with increasing number of psychotic episodes? Is striatal DSS already decreased even before individuals develop prodromal symptoms and enter an at-risk mental state (ARMS), which has been shown to be associated with slightly increased striatal DSS?^64^ This leads to the speculative and possibly controversial question of whether decreased striatal DSS during psychotic remission might represent a “trait” marker linked to increased vulnerability for future psychotic episodes, for example with influences from its baseline level and variability or from previous medication. We believe that these issues are testable and demand further investigation, both longitudinally and at different stages of the disease.

### Unchanged DSS during psychosis

Cross-sectional post hoc tests found no significant differences between patients during psychosis and healthy controls at session 1. Several factors could contribute to this finding: First, we want to stress that medication effects cannot be ruled out completely, despite our statistical control procedures. Second, we interpret our results from the perspective of heterogeneity in schizophrenia across multiple levels, ranging from symptoms to disease course and neurobiological mechanisms. For example, while meta-analyses reported increased striatal DSS in schizophrenia when summarizing results across studies and disease stages,^15,65^ several studies (most of them investigating medicated patients) found no group differences or rather decreased DSS in patients, particularly in those with illness durations comparable to our study (∼12y on average).^19,22–25^ Furthermore, a recent study employing normative modelling of ^18^F-DOPA PET data reported higher levels of extreme deviations in patients, but also a high degree of heterogeneity, suggesting that DSS might be elevated in some but not all patients during psychosis.^66^ Third, heterogeneity within the healthy controls might also play a role. Taken together, our results are generally compatible with the previous literature, but should be interpreted as averages across a spectrum of patients or subgroups, which might impede generalisability.

### Potential relevance of caudate DSS during psychosis for subsequent psychotic relapse

Caudate DSS during psychosis, although on average not changed in patients compared to healthy controls, was significantly higher in patients with subsequent psychotic relapse within 12 months than in patients without relapse (Fig. 4). In contrast, there was no difference regarding caudate DSS during psychotic remission, nucleus accumbens DSS, severity of psychotic symptoms or antipsychotic medication. As an important caveat, these results came from a naturalistic, clinical follow-up with limited sample size and without systematic control of important prognostic factors of psychotic relapse after the second PET measurement, such as medication adherence or substance abuse, and should therefore be viewed as exploratory. Nevertheless, they might provide first insights that higher caudate DSS during a preceding psychotic episode might indicate a more dynamic DSS and thus be relevant for future relapse risk. So far, studies in this field are scarce. Future studies, ideally with three ^18^F-DOPA PET measurements (during psychosis, psychotic remission and psychotic relapse) are needed to clarify the link of our results with: first, studies reporting a relationship between striatal DSS (or related measures), particularly in striatal subregions overlapping with the caudate, during psychotic remission and subsequent psychotic relapse;^28,67^ second, a single-patient report observing increasing striatal DSS from remission to psychotic relapse^27^; and third, other candidates for neurobiological relapse predictors, such as grey matter volume, functional connectivity or gyrification.^68,69^

### Strengths and Limitations

Since longitudinal studies in schizophrenia, particularly those with precise clinical characterisation, are generally rare, we believe that the current work is an important addition to the literature.

There are several limitations in addition to the ones mentioned in the paragraphs above. First, we did not acquire an arterial input function during PET imaging, on which the Patlak model was initially based; instead, we used a cerebellar reference region approach.^70^ However, the widely-used reference region approach appears to be robust also without an arterial input function and can resolve global differences in ^18^F-DOPA uptake.^46,71^ Still, potential differences in cerebellar blood flow between patients and controls could affect our findings.

Second, we did not administer premedication with entacapone and carbidopa, which blocks the formation of peripheral ^18^F-DOPA metabolites, increases the signal-to-noise ratio and is used in many ^18^F-DOPA studies.^20,21,62,72^ Although the exact influence of this premedication on ^18^F-DOPA outcomes and possibly aberrant COMT activity in patients with schizophrenia remains unclear,^73–77^ confounding effects on our results cannot be ruled out completely. However, our approach is consistent with previous ^18^F-DOPA PET studies without premedication,^14,37^ and our reliability analyses demonstrated robustness and comparability of the k_i_^cer^ measures with studies applying premedication, thus strengthening the validity of our measurements.

Third, an inherent issue that could generally confound ^18^F-DOPA PET measurements is that usually, tyrosine hydroxylase is the rate-limiting enzyme in dopamine synthesis, rather than aromatic acid decarboxylase, which metabolizes ^18^F-DOPA.^21,46^

Fourth, all patients took antipsychotic medication, and three patients took mood-stabilising medication. Although we extensively applied statistical control, confounding effects cannot be ruled out completely. Thus, our results should be interpreted with caution and followed up by studies employing stricter medication control or investigating antipsychotic-naïve/free patients.

Fifth, our sample size was modest, but comparable to similar ^18^F-DOPA-PET studies.^21,28,58^

Sixth, the patient group comprised considerably more males than the control group. Although the sex ratio did not significantly differ between patients and controls and no sex effects were found in control analyses, residual effects cannot be ruled out completely.

## Conclusion

First, DSS of both caudate and nucleus accumbens in patients with schizophrenia significantly changed from psychosis to psychotic remission, with a significant longitudinal decrease in caudate. Second, DSS was significantly decreased in patients during early psychotic remission compared to controls in both caudate and nucleus accumbens. Additionally, the exploratory clinical follow-up suggested a link between caudate DSS during psychosis and psychotic relapse within 12 months. Together, these findings demonstrate a link between striatal DSS and the relapsing-remitting course of psychotic symptoms in schizophrenia.

## Data availability

Imaging data were collected in-house and can be provided upon reasonable request.

PET-ki-proc is available on GitHub https://github.com/juliasbrain/PET_ki_proc in the format of Python scripts and can be downloaded as a Docker container. All remaining code used to perform the analyses can be found at https://github.com/juliasbrain/link-that-will-follow.

## Data Availability

All data produced in the present study are available upon reasonable request to the authors.

## Acknowledgements

The authors wish to thank the medical technical assistants Sylvia Schachoff, Amanda Reinhardt and Vivienne Buck of the Department of Nuclear Medicine for their valuable assistance with image acquisition.

