## Supplementary Material for "Striatal dopamine synthesis in schizophrenia decreases from psychosis to psychotic remission"

By Schulz et al.

### $^{18}$F-DOPA PET data processing

Input files for the pet-ki-proc pipeline were dynamic $^{18}$F-DOPA PET images and T1-weighted MPRAGE scans in NifTis format, organised according to the Brain Imaging Data Structure (BIDS) format.[1] The imaging data (measured for 70 min after radiotracer injection) were attenuation-corrected and reconstructed into 30 dynamic frames (1 × 30 s, 10 × 15 s, 3 × 20 s, 2 × 60 s, 2 × 120 s, 12 × 300 s). Motion correction was performed between 5 and 70 minutes post-injection using FSL's *mcflirt* with rigid-body registration, mutual information cost function, and the last frame as reference frame (Supplementary Fig. 1). Denoising was carried out with Chambolle's total variation algorithm (scikit-image, Python), using 1000 iterations and a weight of 100. Then, the atlas-derived regions-of-interest (ROIs) were transformed into subject space using the following steps: (i) each subject's T1w-image was coregistered to the last PET frame using FSL's *flirt* command (correlation ratio cost function, trilinear interpolation); (ii) the resulting image was then spatially normalised to the MNI-152 2 mm standard space; this step involved an affine registration with *flirt*, followed by a non-linear registration with FSL's *fnirt* using the affine matrix from the previous step; (iii) the resulting warp field was inverted using *invwarp* and applied with *applywarp* to bring MNI-152 atlas-derived ROIs into each subject's PET native space. The originally probabilistic ROIs of the striatum (i.e., showing for each voxel its probability of really pertaining to this striatal region) were thresholded at an absolute threshold of 0.6 and binarized, to retain only voxels with high probability of actually belonging to the striatum. Kinetic modeling was performed with Gjedde–Patlak linear graphical analysis,[2] using PET frames between 20 and 60 minutes post-injection and the cerebellum as reference region (probabilistic ROI, i.e., showing for each voxel its probability of belonging to the cerebellum, thresholded at an absolute threshold of 0.9).[3,4] The outcome measure, the influx constant $k_i^{cer}$, was estimated voxel-wise for nucleus accumbens, caudate, and putamen. Mean $k_i^{cer}$ values were then extracted from these ROIs.

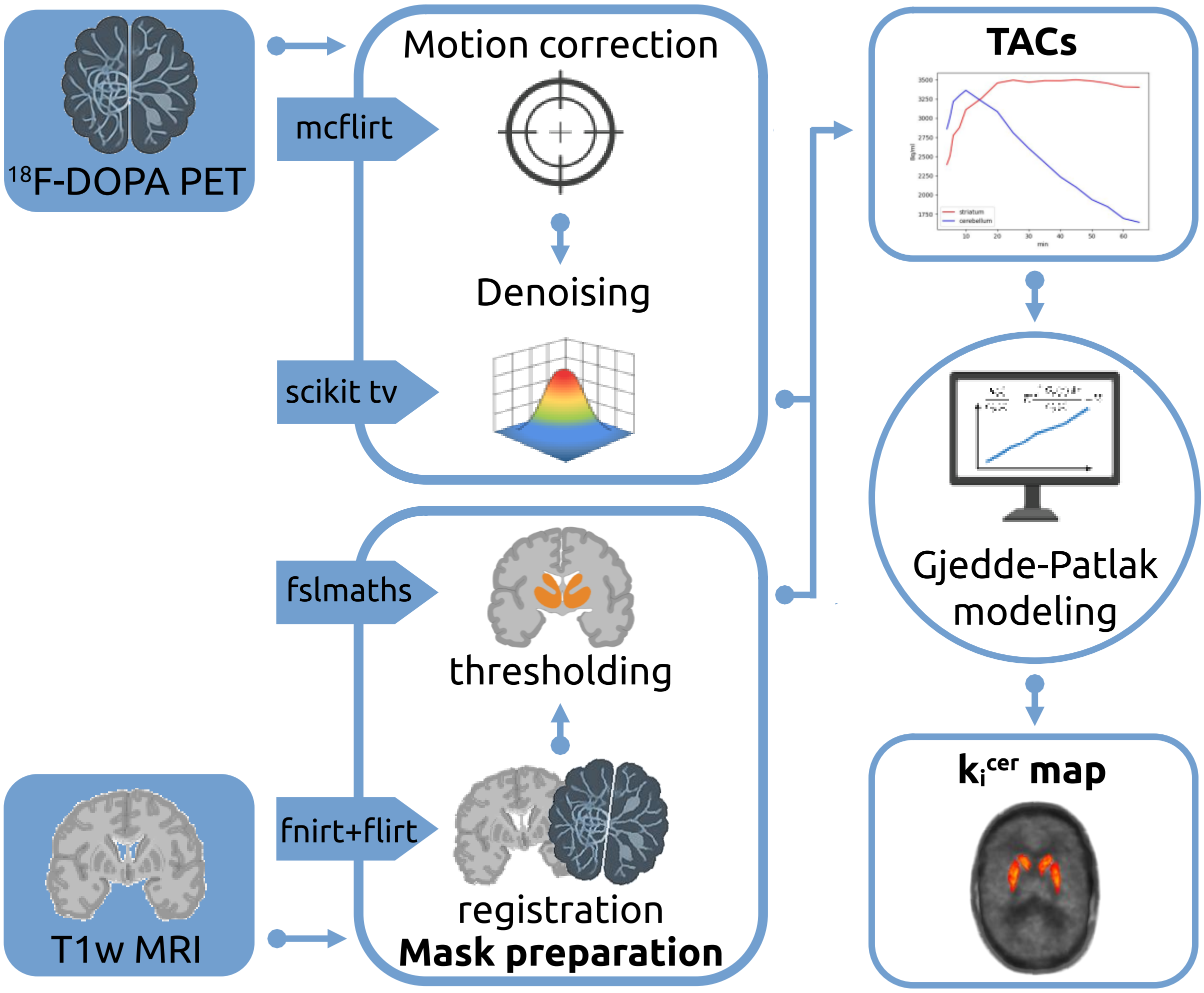

**Supplementary Figure 1 PET processing pipeline.**

For motion correction, dynamic PET frames were aligned to the last frame using FSL's *mcflirt*. Frames were denoised using Chambolle's total variation algorithm implemented in Python's scikit-image library. The T1-weighted anatomical image was coregistered to the native PET space and then normalised to the MNI-152 2 mm standard space using FSL's *flirt* and *fnirt* functions. The resulting warp field was inverted and applied to transform regions of interest (ROIs) into native PET space, where they were thresholded using *fslmaths*. Time-activity curves (TACs) were extracted for each ROI using the cerebellum as reference region. Voxel-wise estimates of the influx constant $k_i^{cer}$ were derived from Gjedde-Patlak linear graphical analysis. Mean $k_i^{cer}$ values were extracted for each striatal subregion to estimate regional dopamine synthesis and storage.

#### Symptom severity and cognition

Similar to PANSS positive, there was a significant decrease in PANSS total and PANSS negative (PANSS-) in patients with schizophrenia from psychosis to psychotic remission (PANSS total ses-01 = 83.71 ± 14.58, ses-02 = 52.87 ± 9.27, $t = -8.8$ $P < 0.001$; PANSS-: ses-01 = 21.89 ± 5.93, ses-02 = 14.96 ± 4.13, $U = 93.5$, $P < 0.001$; Supplementary Fig. 2). Patients showed significantly worse cognitive performance than controls, measured by Trail Making Test part B (TMT-B, linear mixed-effects model main effect of group: $F(1,48) = 23.3$, $P < 0.001$; Supplementary Fig. 3A) and Symbol Coding Task (SCT, linear mixed-effects model main effect of group: $F(1,48) = 81.2$, $P < 0.001$; Supplementary Fig. 3B). Post hoc tests showed worse performance at both time points (TMT-B ses-01: $t = -4.7$, $P < 0.001$, ses-02: $t = -3.7$, $P < 0.001$; SCT ses-01: $t = -4.7$, $P < 0.001$, ses-02: $t = -3.7$, $P < 0.001$). No significant main effect of session (LMM session TMT-B: $F(1,38) = 3.5$, $P = 0.07$; SCT: $F(1,38) = 2.6$, $P = 0.11$), or group x session interaction effect (LMM group x session TMT-B: $F(1,38) = 1.0$, $P = 0.34$; SCT: $F(1,38) = 0.04$, $P = 0.84$) was found for both tests, indicating stable cognitive impairments.

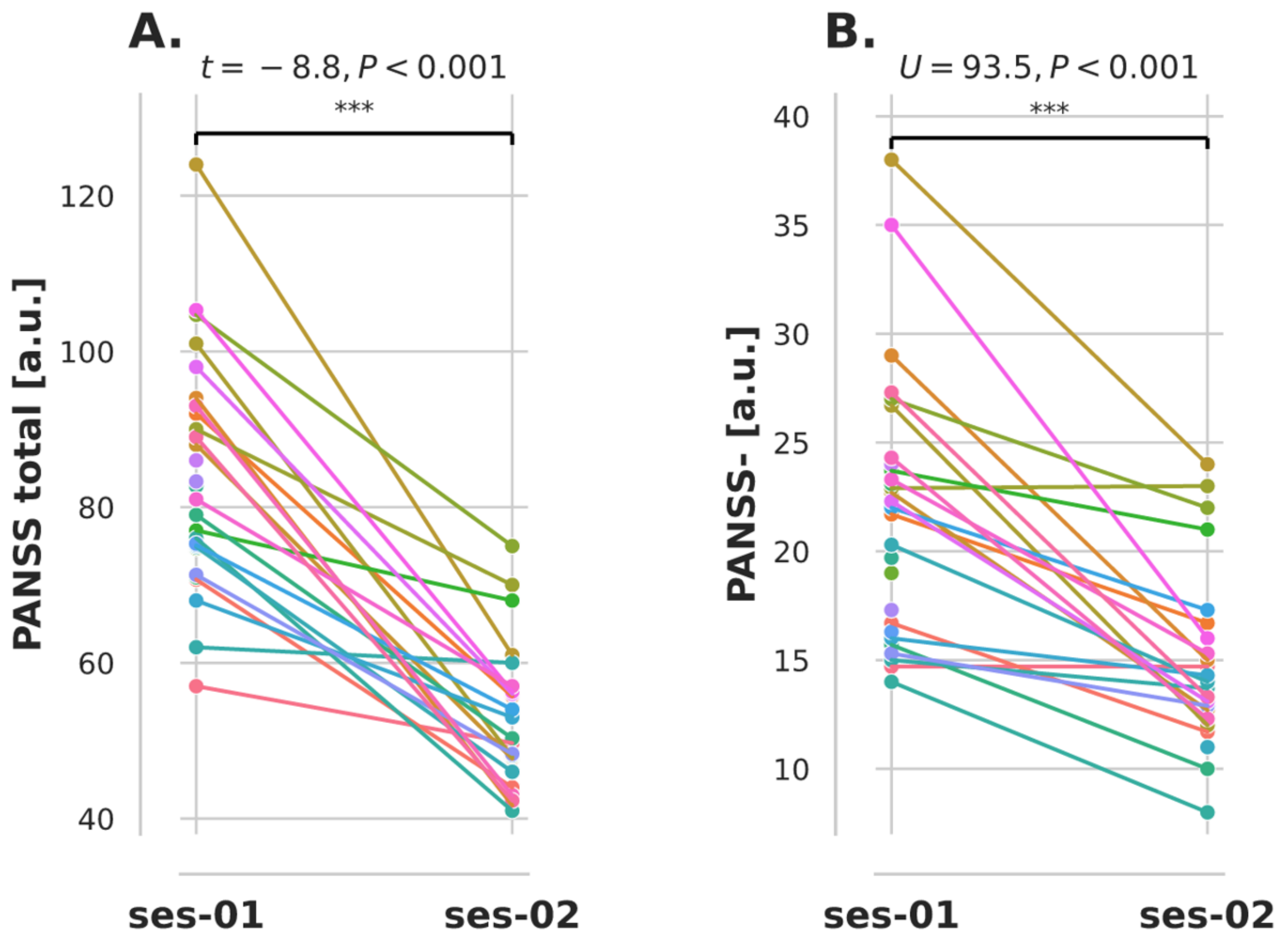

**Supplementary Figure 2 Clinical course of PANSS total and negative.**

Individual trajectories of PANSS total (A) and PANSS negative (B) in patients with schizophrenia from acute psychosis (ses-01) to psychotic remission (ses-02). PANSS total and PANSS negative scores were significantly higher during psychosis compared to remission (PANSS total: $t = -8.8$, $P < 0.001$, PANSS negative: $U = 93.5$, $P < 0.001$). PANSS positive and negative syndrome scale, ses-01 session 1, ses-02 session 2.

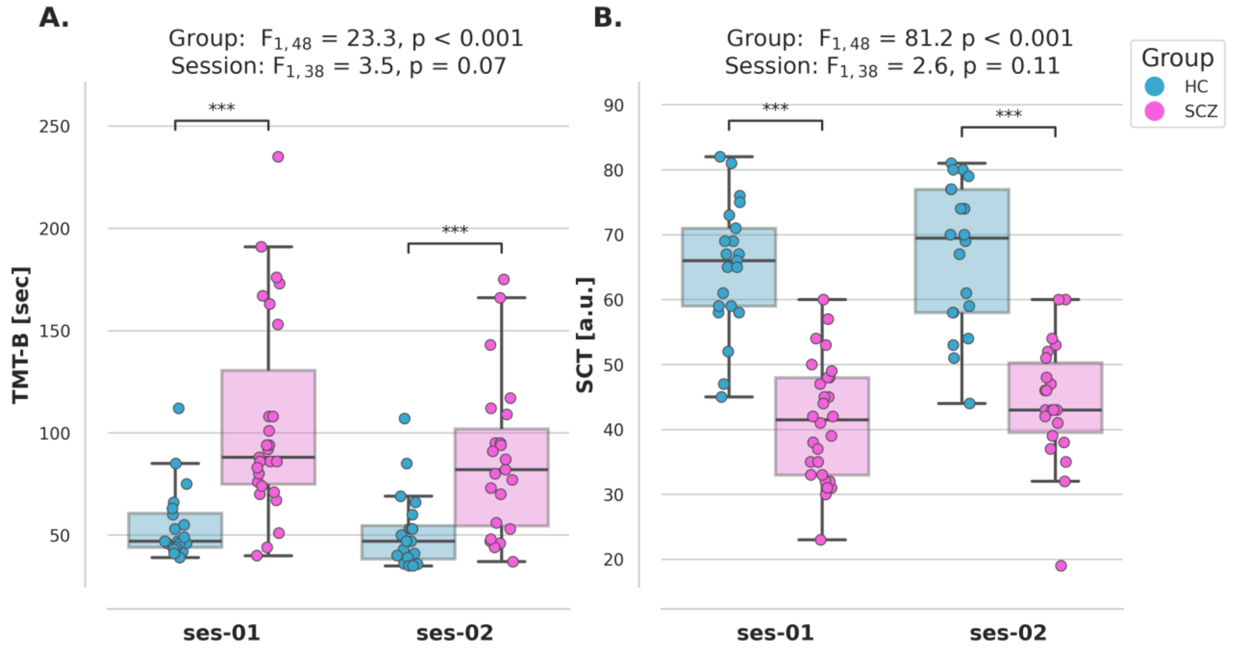

**Supplementary Figure 3 Cognitive performance in patients and controls in each session.** Group comparisons were conducted separately for each cognitive test using linear mixed-effects models. (A) Patients required significantly more time to complete the TMT-B test (LMM group: $F(1,48) = 23.3$, $P < 0.001$), indicating worse performance, in both sessions (post hoc: ses-01: $P < 0.001$; ses-02: $P < 0.001$). There was no main effect of session in both groups (LMM session: $F(1,38) = 3.5$, $P = 0.07$) nor a significant interaction group x session (LMM group x session: $F(1,38) = 1.0$, $P = 0.34$). (B) Patients also achieved significantly lower scores in the SCT (LMM group: $F(1,48) = 81.2$, $P < 0.001$), indicating worse performance, in both sessions (post hoc: ses-01: $P < 0.001$; ses-02: $P < 0.001$). No main effect of session (LMM session: $F(1,38) = 2.6$, $P = 0.11$) or a significant interaction group x session (LMM group x session: $F(1,38) = 0.04$, $P = 0.84$) was found. HC healthy controls, SCT Symbol Coding Test, SCZ patients with schizophrenia, ses-01 session 1 (in patients: psychosis), ses-02 session 2 (in patients: psychotic remission), TMT-B Trail Making Test part B.

### Medication

Oral antipsychotic dose (CPZ equivalents, mean: ses-01 = 470.57 ± 384.74 mg/d, ses-02 = 441.54 ± 311.39 mg/d, $U$ = 322, $P$ = 0.99) and antipsychotic plasma levels (mean: ses-01 = 3.18 ± 3.04, ses-02 = 2.22 ± 1.28, $U$ = 266, $P$ = 0.28) did not differ significantly between sessions in the patient group (Supplementary Fig. 4).

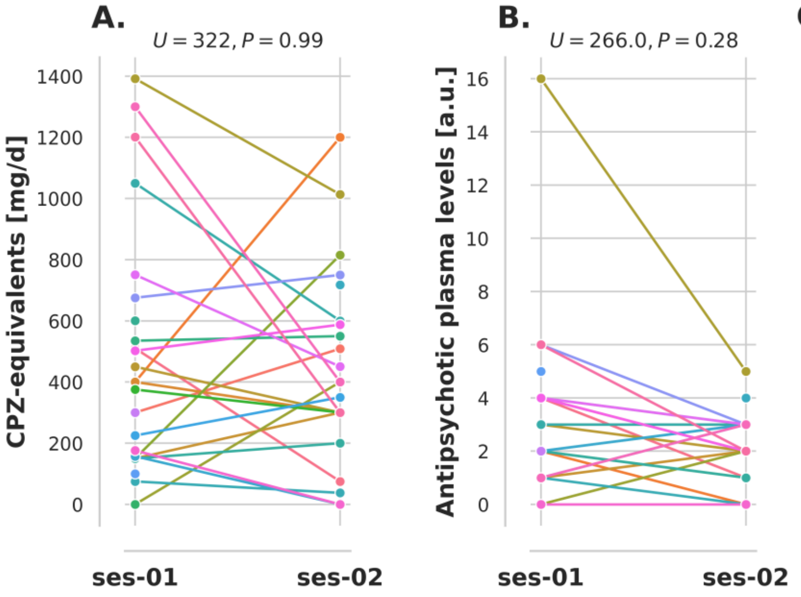

C.

| Antipsychotics | # Patients ses-01 | # Patients ses-02 |
|---|---|---|
| amisulpride | 4 | 4 |
| aripiprazole | 9 | 7 |
| benperidol | 1 | 1 |
| cariprazine | 4 | 1 |
| clozapine | 9 | 8 |
| flupentixol | 1 | 0 |
| haloperidol | 0 | 2 |
| olanzapine | 9 | 6 |
| paliperidone | 1 | 1 |
| pipamperone | 3 | 0 |
| quetiapine | 6 | 4 |
| risperidone | 4 | 0 |
| ziprasidone | 1 | 0 |

**Supplementary Figure 4 Antipsychotic medication.**
Individual trajectories of CPZ equivalents (A) and antipsychotic plasma levels (B) in patients with schizophrenia from acute psychosis (ses-01) to psychotic remission (ses-02). Differences between sessions were evaluated using the Mann-Whitney U test. No significant differences were observed between sessions for either CPZ equivalents ($U$ = 322, $P$ = 0.99) or plasma levels ($U$ = 266, $P$ = 0.28). (C) Number of patients receiving each antipsychotic. CPZ chlorpromazine, ses-01, psychosis, ses-02 psychotic remission.

### Control and reliability analyses

*Reliability analyses of $k_i^{cer}$ measures in healthy controls.* First, we compared regional $k_i^{cer}$ values in healthy controls in the striatum, midbrain and cortical grey matter. To define cortical grey matter, we used the Harvard-Oxford cortical atlas (max. probability threshold 25%), while the midbrain was derived from the CIT168 atlas, combining substantia nigra and ventral tegmental area.[5] In both sessions, the highest $k_i^{cer}$ values were observed in the striatum (ses-01: 0.01320 ± 0.0017 $min^{-1}$, ses-02: 0.01358 ± 0.0018 $min^{-1}$), followed by the midbrain (ses-01: 0.00632 ± 0.0012 $min^{-1}$, ses-02: 0.006 ± 0.0013 $min^{-1}$) and cortical grey matter (ses-01: 0.00171 ± 0.0004 $min^{-1}$, ses-02: 0.00171 ± 0.0003 $min^{-1}$). These results are consistent with the expected hierarchy of dopamine synthesis and storage across these brain regions.[3,6]

Second, we tested for test-retest-reliability of striatal $k_i^{cer}$ measures in healthy controls by calculating intraclass correlation coefficients with two-way mixed-effects models (ICC3k), which quantify the consistency of repeated measurements within the same subjects. They demonstrated excellent test-retest reliability of striatal $k_i^{cer}$ values in caudate and putamen (*ICC3k* = 0.89 and 0.89, respectively) and fair reliability in the nucleus accumbens (*ICC3k* = 0.43). These findings suggest stability of our outcome measure $k_i^{cer}$ in the striatum over time (ratings following reference [7]). Moreover, also the pairwise post hoc test of our principal linear mixed model reported in the main text, comparing HC ses-01 vs. HC ses-02, showed no significant difference in $k_i^{cer}$ between healthy controls' session 1 and session 2 in either nucleus accumbens ($t$ = -1.4, $P$ = 0.48) or caudate ($t$ = -1.4, $P$ = 0.53).

*Effects of imaging-based methodological variables on DSS.* First, we tested for the effect of the reference frame for motion correction by repeating the analysis using the frame at 30 minutes post-injection (i.e., frame No. 23) as reference frame. The group x session interaction remained significant for both nucleus accumbens ($F(1,40)$ = 4.5, $P$ = 0.040) and caudate ($F(1,40)$ = 6.5, $P$ = 0.015).

Second, we assessed the effect of the size of striatum ROIs. In the main analysis, striatum ROIs were thresholded at an absolute value of 0.6. To assess the impact of this parameter, we used alternative thresholds of 0.2, 0.4, and 0.8, and re-calculated our main linear mixed-effects model. Across all thresholds, the group x session interaction remained significant for the caudate and at trend to significance for the nucleus accumbens (Supplementary Table 1).

Third, we evaluated the influence of the size of the cerebellum mask. In the main analysis, the cerebellum ROI was thresholded at 0.9. We repeated the analysis with thresholds of 0.8, 0.85 and 0.95. In all cases, the group x session interaction remained significant (Supplementary Table 1).

| Thresholds | Effects | Nucleus Accumbens | Caudate |
|---|---|---|---|
| **striatal masks** | | | |
| 0.2 | group | $F(1,48) = 2.3$, $P = 0.14$ | $F(1,48) = 1.9$, $P = 0.18$ |
| | session | $F(1,40) = 0.1$, $P = 0.98$ | $F(1,40) = 0.1$, $P = 0.87$ |
| | group x session | $F(1,40) = 3.5$, $P = 0.069$ | **$F(1,40) = 4.9$, $P = 0.032$** |
| 0.4 | group | $F(1,48) = 2.0$, $P = 0.16$ | $F(1,48) = 1.9$, $P = 0.18$ |
| | session | $F(1,40) = 0.1$, $P = 0.99$ | $F(1,40) = 0.1$, $P = 0.92$ |
| | group x session | $F(1,40) = 3.4$, $P = 0.072$ | **$F(1,40) = 5.1$, $P = 0.029$** |
| 0.8 | group | $F(1,48) = 1.0$, $P = 0.33$ | $F(1,48) = 1.7$, $P = 0.20$ |
| | session | $F(1,40) = 0.2$, $P = 0.69$ | $F(1,40) = 0.3$, $P = 0.59$ |
| | group x session | $F(1,40) = 3.5$, $P = 0.067$ | **$F(1,40) = 4.8$, $P = 0.035$** |
| **cerebellar mask** | | | |
| 0.8 | group | $F(1,48) = 2.0$, $P = 0.16$ | $F(1,48) = 1.7$, $P = 0.20$ |
| | session | $F(1,40) = 0.1$, $P = 0.89$ | $F(1,40) = 0.1$, $P = 0.76$ |
| | group x session | **$F(1,40) = 4.4$, $P = 0.041$** | **$F(1,40) = 4.9$, $P = 0.033$** |
| 0.85 | group | $F(1,48) = 2.0$, $P = 0.16$ | $F(1,48) = 1.7$, $P = 0.19$ |
| | session | $F(1,40) = 0.1$, $P = 0.89$ | $F(1,40) = 0.1$, $P = 0.75$ |
| | group x session | **$F(1,40) = 4.5$, $P = 0.040$** | **$F(1,40) = 4.9$, $P = 0.032$** |
| 0.95 | group | $F(1,48) = 2.1$, $P = 0.15$ | $F(1,48) = 1.8$, $P = 0.19$ |
| | session | $F(1,40) = 0.1$, $P = 0.89$ | $F(1,40) = 0.1$, $P = 0.76$ |
| | group x session | **$F(1,40) = 4.6$, $P = 0.038$** | **$F(1,40) = 5.0$, $P = 0.032$** |

**Supplementary Table 1 Results of linear mixed-effects models with different mask thresholds.** The table shows main effects for nucleus accumbens and caudate $k_i^{cer}$ from linear mixed-effects models (dependent variable: mean $k_i^{cer}$; fixed effects: group, session, group × session; random effect: subject) with different thresholds for the striatal and cerebellar masks. Significant effects are indicated in bold.

*Effects of demographic and clinical variables and medication on DSS.* To ensure that the observed alterations in striatal $k_i^{cer}$ were not influenced/confounded by demographic/clinical variables or medication, we conducted control analyses by adding the following covariates to the linear mixed-effects model: (i) age and sex, (ii) smoking status, (iii) between-scan interval, (iv) CPZ equivalents, and (v) plasma levels of antipsychotic medication. In all models, the group x session interaction remained significant for both the nucleus accumbens and the caudate (Supplementary Table 2).

| Covariates | Effects | Nucleus Accumbens | Caudate |
|---|---|---|---|
| i) Age, Sex | group | $F(1,47) = 2.2$, $P = 0.14$ | $F(1,47) = 2.0$, $P = 0.17$ |
| | session | $F(1,39) = 0.1$, $P = 0.85$ | $F(1,39) = 0.1$, $P = 0.71$ |
| | group x session | **$F(1,39) = 4.7$, $P = 0.037$** | **$F(1,39) = 4.9$, $P = 0.033$** |
| ii) Smoking | group | $F(1,47) = 2.1$, $P = 0.16$ | $F(1,47) = 1.7$, $P = 0.20$ |
| | session | $F(1,39) = 0.1$, $P = 0.87$ | $F(1,39) = 0.1$, $P = 0.73$ |
| | group x session | **$F(1,39) = 4.4$, $P = 0.041$** | **$F(1,39) = 4.8$, $P = 0.035$** |
| iii) Between-scan Interval | group | $F(1,47) = 2.0$, $P = 0.16$ | $F(1,47) = 1.7$, $P = 0.20$ |
| | session | $F(1,39) = 0.1$, $P = 0.87$ | $F(1,39) = 0.1$, $P = 0.73$ |
| | group x session | **$F(1,39) = 4.1$, $P = 0.049$** | **$F(1,39) = 5.0$, $P = 0.031$** |
| iv) CPZ-equivalents | group | $F(1,47) = 2.1$, $P = 0.16$ | $F(1,47) = 1.7$, $P = 0.20$ |
| | session | $F(1,39) = 0.1$, $P = 0.88$ | $F(1,39) = 0.3$, $P = 0.59$ |
| | group x session | **$F(1,39) = 5.2$, $P = 0.028$** | **$F(1,39) = 5.1$, $P = 0.030$** |
| v) Antipsychotic Plasma Levels | group | $F(1,47) = 2.0$, $P = 0.16$ | $F(1,47) = 1.7$, $P = 0.20$ |
| | session | $F(1,39) = 0.1$, $P = 0.87$ | $F(1,39) = 0.1$, $P = 0.73$ |
| | group x session | **$F(1,39) = 4.1$, $P = 0.049$** | **$F(1,39) = 5.1$, $P = 0.030$** |

**Supplementary Table 2 Results of linear mixed-effects models with covariates.**
The table shows main effects for nucleus accumbens and caudate $k_i^{cer}$ from linear mixed-effects models (dependent variable: mean $k_i^{cer}$; fixed effects: group, session, group × session; random effect: subject) with different covariates. Significant effects are indicated in bold. CPZ chlorpromazine.

*Effects of specific antipsychotic drugs on DSS.* To further investigate the effect of specific antipsychotic drugs on the results, we selected three antipsychotics of particular interest for control analyses: clozapine, which might particularly influence DSS,[8] aripiprazole, which is a partial dopamine receptor agonist and might influence DSS,[9] and amisulpride, which is a rather selective D2 antagonist and could affect dopamine transmission in a particular way.[10,11] For each antipsychotic, we conducted control analyses by adding a binary covariate (1 if the patient was taking the drug, 0 if not) to the linear mixed-effects model. The group x session interaction remained significant in all cases: clozapine, $P = 0.017$ for caudate and $P = 0.041$ for nucleus accumbens; aripiprazole, $P = 0.016$ for caudate and $P = 0.043$ for nucleus accumbens; amisulpride, $P = 0.013$ for caudate and $P = 0.042$ for nucleus accumbens. This suggests no substantial influence of these drugs on the results.

### Exploratory follow-up: ‘Clinical’ differences between relapsing and non-relapsing patients

At the clinical follow-up 12 months after the measurement in psychotic remission, oral antipsychotic dose (CPZ equivalents) was higher in patients who had had a psychotic relapse compared to those who had not ($t = -2.4$, $P = 0.03$); plasma levels were not measured at follow-up. Increased oral doses of antipsychotic medication after relapsing psychosis might reflect the clinical treatment of affected vs. non-affected patients.

Concerning ‘clinical’ differences between relapsing and non-relapsing patients in the stages preceding the relapse, we compared medication, severity of psychotic symptoms and between-scan interval (as potential hint at the recovery time of psychotic symptoms) during the stages of psychosis and psychotic remission before the relapse between patients with and without subsequent psychotic relapse. Neither during psychosis nor during psychotic remission did we observe any differences in CPZ equivalents (ses-01: $U = 56.5$, $P = 0.81$; ses-02: $t = -0.5$, $P = 0.60$), antipsychotic plasma levels (ses-01: $U = 61.0$, $P = 0.57$; ses-02: $t = 1.5$, $P = 0.15$) or severity of psychotic symptoms (PANSS+; ses-01: $t = -0.03$, $P = 0.98$; ses-02: $t = -1.2$, $P = 0.23$). However, patients with subsequent psychotic relapse had a significantly longer between-scan interval ($t = 3.4$, $P = 0.003$).
